# Validation of the self-test computerised PROTECT Cognitive Test System for detection of dementia and Mild Cognitive Impairment

**DOI:** 10.64898/2026.07.23.26358814

**Authors:** Anne Corbett, Kim Idar Giske, Abbie Palmer, Millie Sander, Christine Davis, Kate Stych, Mary O’Leary, Vincent Hayman, Adam Bloomfield, Jon Huntley, Dag Aarsland, Nicolas Castellanos-Perilla, Felipe Botero-Rodriguez, Gerard Griffioen, Mieke Nuytten, Chris Fox, Louise Allan, David Llewellyn, Nicholas Ashton, Hanna Huber, Adam Hampshire, Jeffrey Cummings, Sallie Lamb, Jane Atwell-Thomas, Clive Ballard

## Abstract

Detection and characterisation of dementia and Mild Cognitive Impairment (MCI) is essential for diagnosis and to support recruitment of patients into trials of disease-targeted therapies. Computerised systems offer a means of improving detection in community and primary care settings in a scalable way. This study presents further validation of the self-test PROTECT Cognitive Test System of eight assessments of memory, attention and executive function in 36,941 participants (35,822 healthy, 1046 MCI, 73 dementia). PROTECT shows robust separation of dementia and non-dementia participants (*p* < 0.001), and good discriminative ability for dementia (Area Under the Curve = 0.966) with 90.90% sensitivity and 87.80% specificity. An optimised detection algorithm robustly identified MCI (P<0.001) and predicted 24-month decline across cognitive domains in both amnestic MCI and non-amnestic MCI phenotypes compared with healthy controls. PROTECT cognitive data also correlated strongly with the plasma biomarkers p-tau217 and Neurofilament Light (*n* = 46). The system provides a means of improving dementia and MCI detection using self-testing, offering a scalable triage and monitoring tool for clinical pathways and trials.

## Introduction

There are one million people with dementia in the UK, of which Alzheimer’s Disease (AD) is the most common cause, generating an annual cost of £42billion^1^. Mild Cognitive Impairment (MCI) affects up to 15% of adults over 50 with 10% developing dementia annually^2^. Despite the importance of diagnosis and support, many people with dementia and MCI are not identified and may wait until they reach a crisis before seeking help^3^. Emerging disease-targeted therapies have the potential to change the landscape of AD treatment but will be most effective in people with early and pre-clinical impairment. This highlights the need for an efficient patient funnel in the community and primary care to support referral and diagnosis into specialist settings^4-6^, as well as to facilitate large-scale recruitment of preclinical groups into clinical trials. Existing health services models do not have capacity or suitable assessment tools to support large-scale patient triage for diagnosis or trials and there is a need to integrate scalable approaches, using self-test methodologies, to improve capacity into these pathways.

A further important issue relates to the complexity presented by MCI. Individuals present with differential trajectories and patterns of impairment, with single and multi-domain impairment across amnestic and non-amnestic domains. Fluctuating and reverting MCI is also common, with 20% of patients reverting to healthy cognition during followup. Amnestic MCI is a common prodromal state for AD whereas non-amnestic MCI may potentially be indicative of broader range of trajectories, including non-Alzheimer dementia. It will be critical to improve detection of these important patient groups to improve the pre-clinical patient funnel and enable earlier diagnoses.

Diagnosis of dementia currently involves a combination of clinical history taking, neuropsychological testing, neuroimaging and, increasingly, blood biomarker assessment^7^. This process is intensive, invasive and expensive and requires efficient triage to create an effective diagnostic pathway. Traditional paper and pencil assessments of cognition, such as the Mini-Mental State Examination (MMSE), Addenbrooke’s Cognitive Assessment (ACE), Alzheimer’s Disease Assessment Scale-Cognitive Subscale (ADAS-Cog) and the Montreal Cognitive Assessment (MoCA) have learning effects, have limited ability to detect changes in specific cognitive domains, and rely on a trained health professional^8^. As a result, these tools are not suitable for scalable cognitive assessment.

Computerised cognitive testing and remote assessment offers a means to improve on the current status quo for brain health assessment for both clinical and research applications. Computerised tests offer consistent, unbiased, highly sensitive recording of cognitive function, which is key to diagnosis and early detection of cognitive decline^9-12^ as well as for quantifying treatment outcomes in clinical trials^13^. Importantly, computerised tests raise the potential for remote, unsupervised assessment through self-testing which makes them scalable^14^ for deployment in the community and in primary care without adding burden to physicians. Over 30 digital cognitive test systems exist, and the field is rapidly growing^15^. However, the majority of computerised cognitive test systems have been used predominantly in supervised clinical settings, usually for clinical trial ^10,16^⍰. There is a clear opportunity to position computerised testing as a means to scale the clnical diagnostic pathway through self-testing, with patients using computeri^1414^⍰. Preliminary self-test validation is available for several computerised systems, but the largest validation study is the PROTECT-UK cohort which uses a computerised test system embedded in the study website.

The PROTECT Cognitive Test System is a suite of computerised neuropsychological assessments that has been developed and validated as part of a portfolio of research over the last ten years. It has undergone full computer systems and data validation in the PROTECT-UK cohort of 50,000 participants, with over 2.5 million self-test assessments completed^17^. The system has shown ability to detect statistically meaningful age-related cognitive decline, demonstrating concurrent validity with established Food and Drug Administration (FDA)-approved criteria for early cognitive impairment, and sensitivity to trajectory of cognitive decline in healthy and impaired individuals^18^. It has been used to measure cognitive change in several completed and ongoing clinical trials and ageing cohorts, which provide the opportunity to further validate its sensitivity, including definiting age-sensitive normative data and thresholds, to establish its accuracy in detecting clinically-meaningful cognitive status and change^19-23^.

There is also an opportunity to combine computerised neuropsychology with rapidly emerging blood biomarkers for neurodegeneration. Blood biomarkers are becoming well established for AD risk detection and are now being adopted into clinical practice^24^. The most robust diagnostic blood biomarker is phosphorylated tau at amino acid 217 (p-tau217), which shows highest accuracy for detection of amyloid pathology ^25,26 27,28^. Neurofilament light chain (NfL) is also a commonly cited biomarker of neurodegeneration indicating axonal and neuronal damage^29^. The combination of blood biomarkers with self-test computerised neuropsychology offers the potential for scalable diagnosis, monitoring, and assessment to support clinical diagnostic pathways through primary care and to specialist services or clinical trials.

This study builds upon the current validation of the self-test PROTECT cognitive system, reporting data from clinical dementia cohorts and the PROTECT-UK longitudinal cohort study of more than 35,000 participants. It aimed to establish the sensitivity and specificity of the system for dementia and MCI, to define age-sensitive thresholds to enable flagging of AACD, MCI and dementia based on computerised self-testing at home, and to validate them both cross-sectionally and longitudinally in a large dataset of healthy, MCI and dementia patients. The study also aimed to establish whether the PROTECT system correlates with the p-tau217 and NfL blood biomarkers.

## Methods

### Study Design

This was a longitudinal data analysis study using data from the self-test PROTECT Cognitive Test System that sought to establish and validate age-sensitive thresholds for MCI and dementia and to evaluate their predictive validity for cognitive change.

### Source Cohorts

The study used data collected in the national PROTECT-UK online ageing cohort which received ethical approval from the London Bridge National Health Service Research Ethics Committee (13/LO/1578) and launched in April 2014. This study analysed data collected between April 2014 and January 2026. A second comparator dataset was derived from patients with dementia in five international sites in the UK, Spain, Norway and the Netherlands^13,16,17,20,21^ as a comparator dataset for dementia sensitivity analyses. Ethical approval for this dementia sub-group data collection was obtained from the (NorthWest Greater Manchester East NHS Research Ethics Committee Ref: 24/NW/0151).

### Participants

All participants were aged between 50 and 90 years old, had capacity to consent and were able to use a computer or touchscreen device. PROTECT-UK participants were healthy volunteers residing in the UK with no self-reported diagnosis of dementia. Participants were excluded if they had a visual impairment, non-dementia neurological condition, stroke affecting upper limbs or any physical disability impairing the use of a device. AD patients fulfilled criteria for mild to moderate AD based on site-specific diagnostic protocols and fulfilment of the National Institute of Neurological and Communicative Disorders and Stroke and Alzheimer’s Disease and Related Disorders Association (NINCDS-ADRDA) criteria with impairment in at least two cognitive domains with impact on functionality.

### Recruitment and consent

Participants with dementia were recruited from clinical sites in the UK, Spain, Norway and the Netherlands^13,16,17,20,21^. Patients were pre-screened for eligibility criteria and provided written informed consent. Cognitively healthy participants in the PROTECT-study were recruited across the UK as part of the study open recruitment phase between 2014 and 2024. Registration and informed consent were performed online through an ethically approved digital process, including consent for use of data for research purposes. All participants provided demographic information and personal information at registration.

### Cognitive Assessments

Participants completed the PROTECT Cognitive Test System independently on a computer or touchscreen device. The system consists of eight individual tests for working memory, episodic memory, executive function and attention. The tests are Paired Associate Learning, Digit Span, Self-Ordered Search, Simple Reaction Time, Choice Reaction Time, Digit Vigilance, Delayed Picture Recognition and Verbal Reasoning. Description of these tests and their outputs are shown in Supplementary Table 1. Participants completed a brief practice session of each test in sequence prior to completing a full test session to reduce practice effects due to orientation to the system. Tests run in a pre-scheduled sequence and start automatically to ensure consistency of delivery. Participants in the PROTECT-UK cohort completed the Cognitive Test System annually as part of the longitudinal data collection protocol. Participants accessed the system from the PROTECT-UK study online dashboard and completed the tests unsupervised at home. Participants in the AD cohort completed the test system at a clinical site. Staff were available to support in the event of a technical or medical issue but did not assist in completion of the tests themselves.

### Blood Biomarker Collection

A subset of participants provided a venous blood sample by venepuncture for biomarker analysis at a clinic visit, taken by a trained staff member. Prior to analysis, plasma samples were thawed, vortexed for 30 seconds at 2,000 rpm and centrifuged at 4,000 g and 20°C for 10 minutes. Biomarker analysis was completed by Single molecule array (Simoa) technology using a 2X dilution protocol according to manufacturer’s instructions (ALZpath Simoa^®^ p-tau217 and NfL v2 Assay,) (Quanterix, Billerica, MA, USA) to detect calibrated levels of p-tau217 and NfL.

### Data handling

Data were transformed to a normal distribution using a *log1p* (log ^30^) function to compress the tail and approximate a normal distribution and outliers removed using a vectorised approach (Figure 1). This is essential for self-test data to filter out invalid test cases, for example, if a participant abandons a test mid-way through, without deleting genuine clinical decline data.

**Figure 1:**
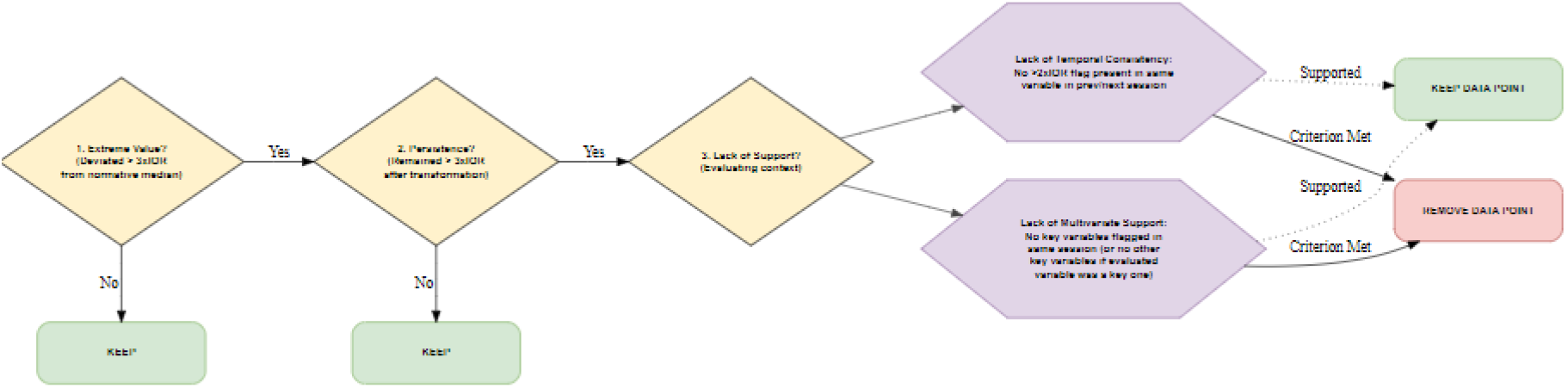
High performance vectorised filtration algorithm logic for outlier removal.

**Figure 1:**
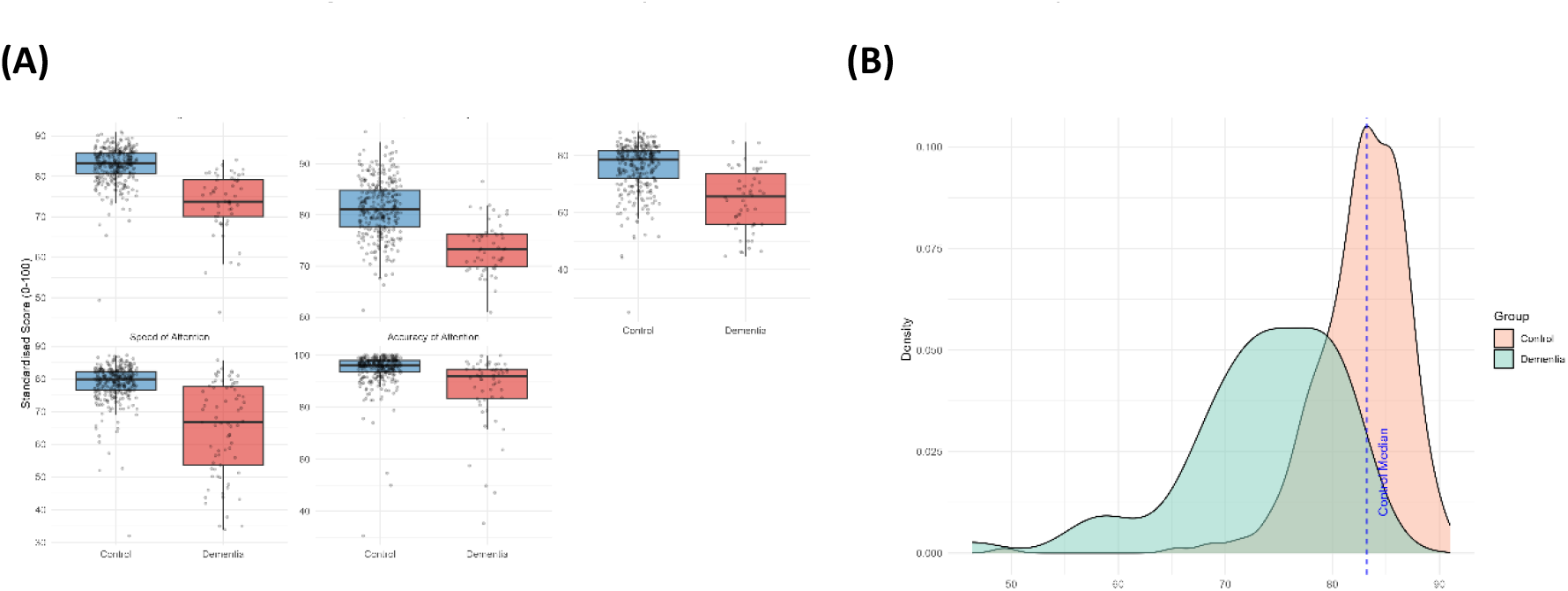
The PROTECT Cognitive Test System detects differences in performance in all cognitive domains between age-matched healthy controls and dementia patients.

Four primary cognitive domain composites - Memory, Speed of Attention, Accuracy of Attention, and Executive Function – and a combined global composite were calculated and standardised to a uniform 0-100 scale. Strict data completeness thresholds were enforced to mitigate the impact of missing data while preserving construct validity (Supplementary Methods XX). Normative thresholds were calculated using five-year age bands: 50-54, 55-59, 60-64, 65-69, 70-74, 75-79, and 80+. Mean and standard deviation were calculated and converted back into specific raw-score values, and thresholds were generated for Age-Associated Cognitive Decline (AACD), MCI and dementia based on the published FDA criteria of 1, 1.5 and 2SD respectively.

### Validation for dementia detection and correlation with AD biomarkers

A 1:4 age- and sex-matched healthy comparator dataset was derived from the PROTECT-UK cohort to evaluate sensitivity to dementia. Independent paired t-test analysis compared cognitive performance on all variables in the PROTECT-UK cohort and dementia cohort, with Bonferroni correction applied for multiple comparisons. Diagnostic utility was assessed via Receiver Operating Characteristic (ROC) curve analysis, reporting the Area Under the Curve (AUC), sensitivity and specificity to dementia. Variables across the five primary cognitive domains were selected to establish an algorithm for detection of dementia and MCI. Variable selection prioritised tests that maximised sensitivity to dementia in the confirmatory cohort whilst maintaining robust specificity against the normative baseline cohort. Concurrent validity with blood biomarker levels was analysed using Spearman’s rank correlation.

### Validation for MCI detection and longitudinal decline

The MCI thresholds were applied to the dataset for longitudinal validation. Individuals fulfilling criteria for MCI at baseline, but showing reverting or fluctuating phenotypes back to normal cognition were excluded based on 48-month trajectory. The remaining confirmed MCI cases were classified as either amnestic MCI (impairment in at least one memory domain) or non-amnestic MCI (impairment in attentional and/or executive domains but not memory). 24-month longitudinal trajectories for aMCI and non-aMCI were analysed using Linear Mixed-Effects Models (LMM) fit via restricted maximum likelihood (REML) to account for missing data and intra-individual correlation over repeated measures. To ensure the observed subtype trajectories represented genuine clinical phenotypes rather than statistical artifacts of the MCI flagging logic, the groups were evaluated against an independent variable from each domain that was not used to classify the MCI status. Through an exploratory *ad hoc* analysis utilising the same 24-month LMM structure, we examined whether the baseline-defined subtypes exhibited distinct longitudinal progression patterns on these untrained domains. All analyses were completed in R Studio (Version 4.5.0).

## Results

### Cohort Characterisation

The total baseline cohort from PROTECT-UK comprised 43,974 participants. Following exclusions for reverting/fluctuating trajectories and missingness, the final analytical baseline cohort comprised 35,822 controls and 1,046 MCI cases. The dementia comparator cohort comprised 73 participants, and the age-matched cohort contained 316 participants. Baseline demographics were consistent across the PROTECT-UK subgroups and the dementia comparator groups (Table 1). Blood biomarker data were available from 46 participants across the healthy (n = 19) and dementia (n = 27) cohorts.

**Table 1:** Cohort demographics.

| Group | Baseline (n) | Age<br>Mean (SD) | Female (%) |
| --- | --- | --- | --- |
| **Cognitively Healthy** | 35,822 | 62.5 (7.6) | 71.4 |
| **Amnesic MCI** | 378 | 65.0 (8.7) | 69.3 |
| **Non-Amnesic MCI** | 668 | 63.4 (7.6) | 73.8 |

| Dementia comparator cohorts |  |  |  |
| --- | --- | --- | --- |
| **Dementia cohort** | 73 | 73.77 (8.33) | 39.0 |
| **Age-matched healthy cohort**<br>**(derived from non-MCI group)** | 316 | 73.03 (6.98) | 39.0 |

### Differentiation between dementia and healthy participants

Comparative analysis of cognitive performance between the dementia and age-matched healthy cohort showed significant difference between all cognitive test outputs and composite measures for global cognition, memory, speed of attention, attentional accuracy and executive function (P<0.001) (Supplementary Table 1, Figure 1).

### Selection of variables for detection of dementia and MCI

A ROC analysis using the comparator dataset of participants with dementia identified five optimal variables for the flagging algorithm with AUC of >0.8 for all variables (Table 2, Figure 3). Key variables for memory and executive function showed robust sensitivity to dementia (Executive function: sensitivity 90.7%, specificity 78.7%; Memory: sensitivity 90.9%, specificity 82.10%). A global cognition composite measure showed sensitivity of 90.9% and specificity of 87.8% and AUC of 0.966. Independent validation variables with the next best sensitivity were selected from the remaining variables except for executive function, for which there was no available variable (Supplementary Table 1)

**Table 2:** Sensitivity and specificity of optimal test system variables for detection of dementia.

| Cognitive Domain | Variable | AUC | 95% CI | P |
| --- | --- | --- | --- | --- |
| Executive Function | Verbal Reasoning Total Score | 0.921 | 0.887 - 0.955 | <0.001 |
|  | Composite | 0.880 | 0.833 - 0.927 | <0.001 |
| Memory | Digit Span Total | 0.882 | 0.845 - 0.920 | <0.001 |
|  | Picture Recognition Accuracy | 0.865 | 0.825 - 0.904 | <0.001 |
|  | Composite | 0.935 | 0.909 - 0.960 | <0.001 |
| Speed of Attention | Choice Reaction Time Speed | 0.862 | 0.813 - 0.912 | <0.001 |
|  | Composite | 0.855 | 0.806 - 0.905 | <0.001 |
| Accuracy of Attention | Digit Vigilance Accuracy | 0.873 | 0.832 - 0.913 | <0.001 |
|  | Composite | 0.837 | 0.784 - 0.890 | <0.001 |
| **Global Cognition** | Composite | 0.966 | 0.941 - 0.972 | <0.001 |

**Figure 3:**
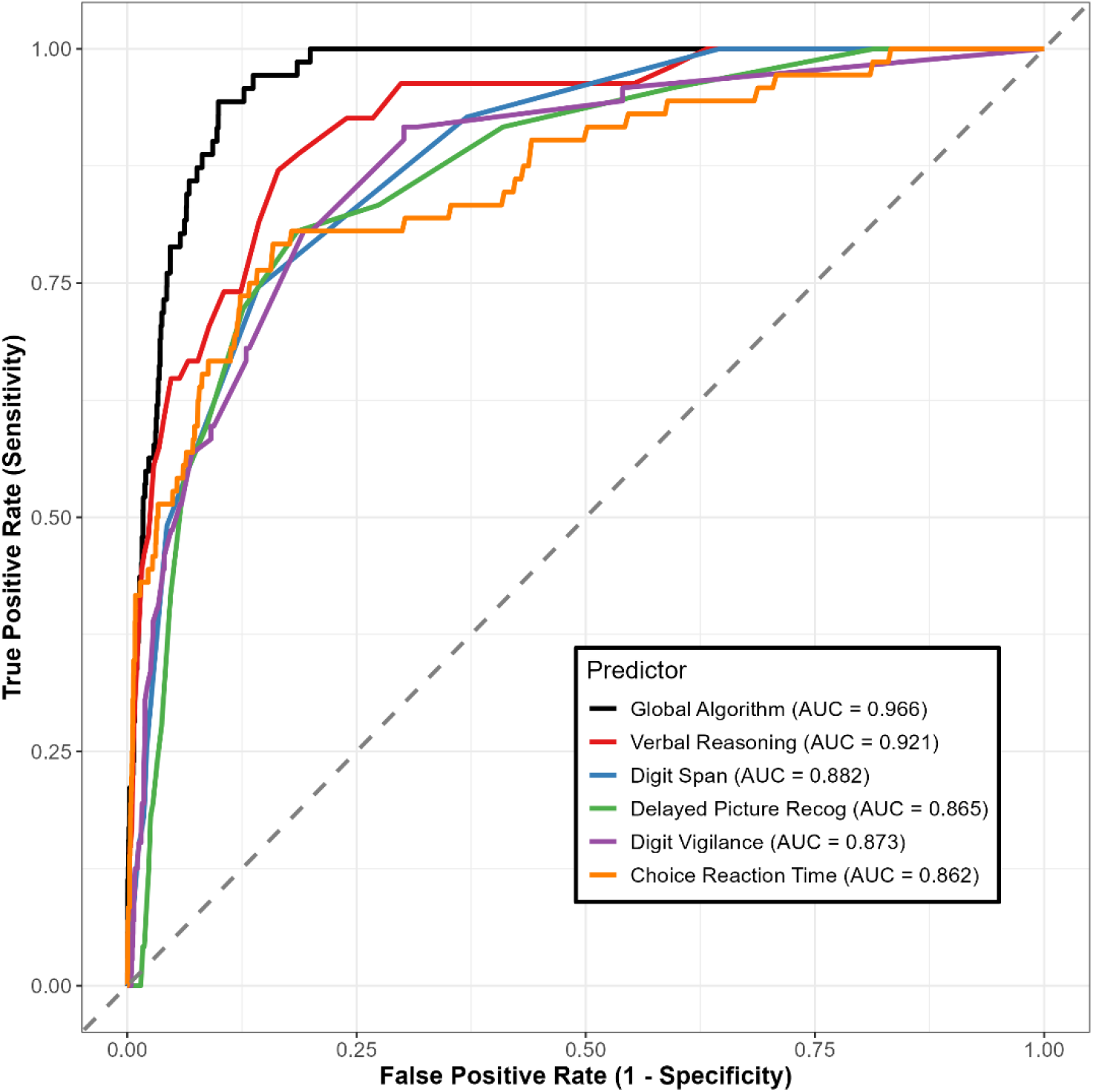
Receiver Operating Characteristic (ROC) curves demonstrating diagnostic discriminative accuracy of the global dementia/MCI predictor algorithm and PROTECT system composite measures.

### Correlation with venous blood biomarkers for neurodegeneration

Analysis showed significant correlations between p-tau217 and all cognitive domains (P<0.001) including global cognition (P=0.003). NfL showed significant correlation with memory, speed of attention, accuracy of attention and executive function domains but not with global cognition (Table 3).

**Table 3:** Correlation of the PROTECT Cognitive Test System with blood biomarkers p-tau217 and NfL (n=46)

| Cognitive Domain | Plasma p-tau217 |  | Plasma NfL |  |
| --- | --- | --- | --- | --- |
| | Spearman's $\rho$ | P Value | Spearman's $\rho$ | P Value |
| Global Cognition | 0.438 | 0.003 | 0.13 | 0.405 |
| Memory Composite | 0.707 | <0.001 | 0.547 | <0.001 |
| Speed Composite | 0.561 | <0.001 | 0.49 | 0.001 |
| Accuracy Composite | 0.661 | <0.001 | 0.442 | 0.004 |
| Executive Function Composite | 0.604 | <0.001 | 0.416 | 0.011 |

### Detection of MCI at baseline

At baseline the amnestic and non-amnestic MCI subgroups performed significantly worse than the non-MCI cohort in all variables and composite measures including the independent variables (P<0.001) (Table 4).

**Table 4:** Cross-sectional comparison of cognitive performance in healthy and MCI participants defined by the MCI prediction algorithm.

| Domain | Amnesic MCI |  |  | Non-Amnesic MCI |  |  |
| --- | --- | --- | --- | --- | --- | --- |
|  | Baseline Mean (Z-Score) | Mean Difference (vs Non-MCI) (Z-Score) | Adj. P (FDR) | Baseline Mean (Z-Score) | Mean Difference (vs Non-MCI) (Z-Score) | Adj. P (FDR) |
| Global Cognition | -2.17 | -2.06 | < 0.001 *** | -2.04 | -1.95 | < 0.001 *** |
| Memory | -1.96 | -1.85 | < 0.001 *** | -0.73 | -0.66 | < 0.001 *** |
| Executive Function | -1.29 | -1.18 | < 0.001 *** | -1.98 | -1.87 | < 0.001 *** |
| Accuracy of Attention | -1.20 | -1.17 | < 0.001 *** | -1.65 | -1.63 | < 0.001 *** |
| Speed of Attention | -1.08 | -1.03 | < 0.001 *** | -1.44 | -1.40 | < 0.001 *** |

Cross-domain profiling showed differential profiles for the amnestic and non-amnestic groups where amnestic MCI is characterised by a dominant impairment in episodic memory whilst the non-amnestic MCI group showed greatest impairment in attentional accuracy (Figure 4).

**Figure 4:**
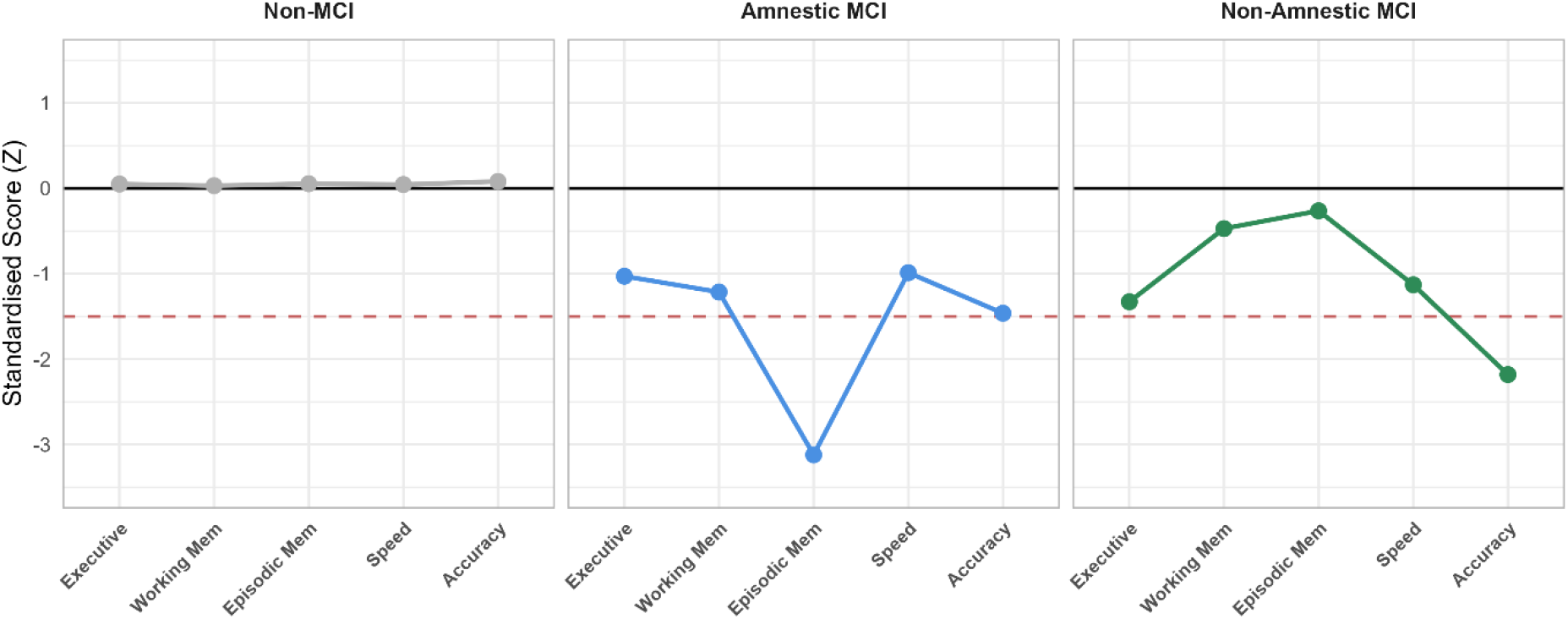
MCI Subtype Cohorts show differential performance across cognitive domains.

### Prediction of decline in amnestic and non-amnestic MCI

The clinical meaningfulness and prognostic value of the MCI detection algorithm was validated for cognitive trajectory over 24 months, with a primary research question examining the predictive power of for amnestic impairment, given the relevancy of amnestic impairment for early AD detection. The amnestic MCI group showed significant decline in memory (P=0.034) and executive function (P<0.001) composites, as well as in the independent validation variables for memory (P<0.001). The non-amnestic MCI group showed significant decline across all composite measures (P<0.001) and validation variables except for one accuracy of attention variable (Figure 5, Table 5).

**Table 5:**
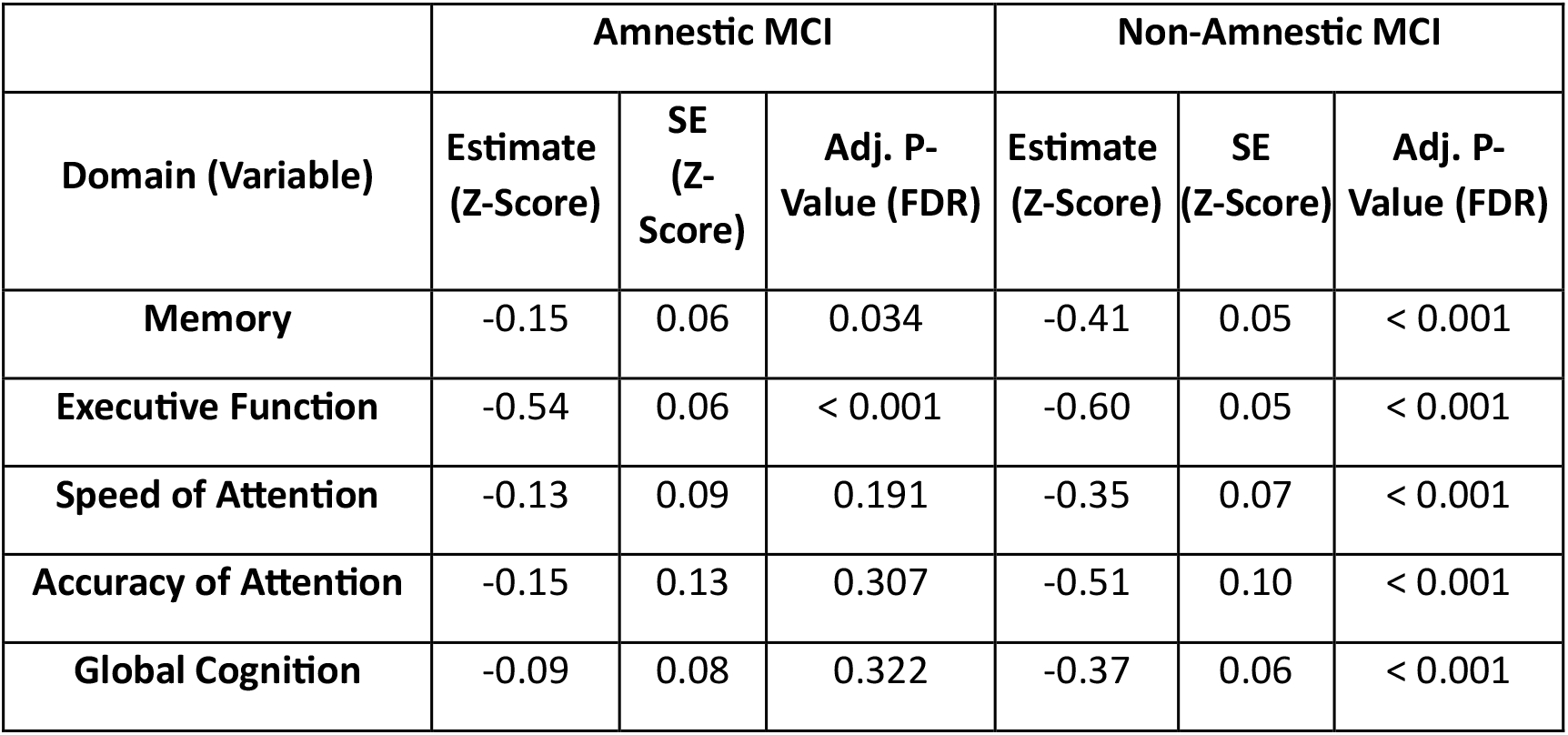
Change in cognition over 24 months in MCI group vs non-MCI group.

**Figure 5:**
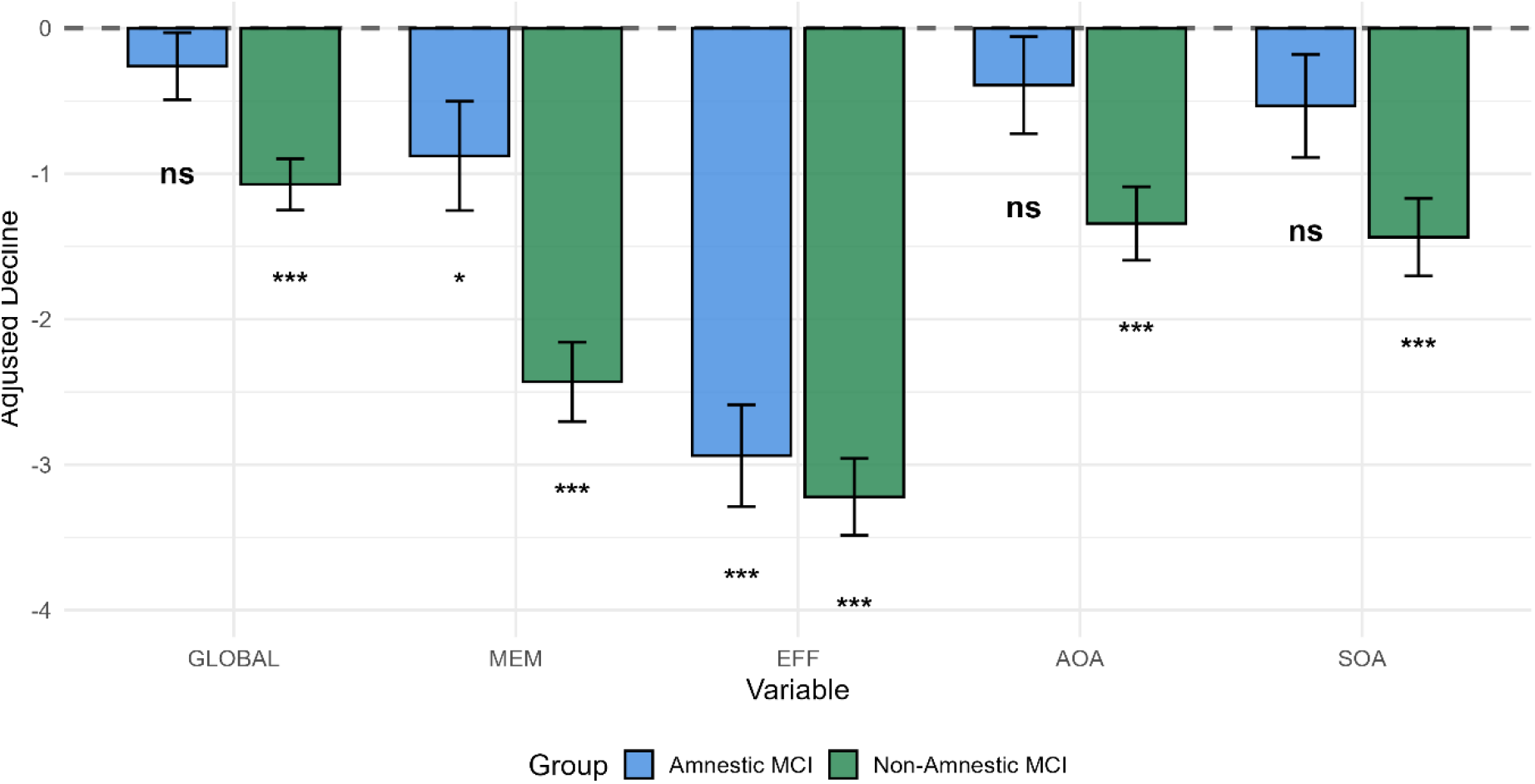
24-month longitudinal cognitive decline in amnestic and non-amnestic MCI relative to healthy controls.

## Discussion

This study expands the validation of the PROTECT self-test computerised Cognitive Test System in a cohort of more than 35,000 participants and demonstrates an improved thresholding approach to identify dementia and MCI using the test system. The outcomes show that this simple to use, self-test system has excellent sensitivity and specificity for dementia and robust predictive validity for cognitive decline. It also has excellent concurrent validity with the well validated plasma tau217 AD biomarker and with the plasma NfL neurodegeneration biomarker, raising the opportunity to combine these two measures to improve early detection and triage of pre-clinical groups.

The comparison of performance between dementia and healthy cohorts shows that the PROTECT system identifies significant differences in performance in all tests across all cognitive domains of memory, attention, speed and executive function. This is further strengthened by the ROC analysis which shows good discriminative ability across all domains with highly significant AUC values. For global cognition this reaches 0.966, showing sensitivity to dementia of 90.9% and specificity of 87.8%. This meets the generally accepted thresholds for clinical utility^31^ and compares favourably with existing digital cognitive assessment devices such as CANTAB Mobile and CogState, which have reported AUC of 0.8-0.95, dementia sensitivities of 80-97% and specificities of 70-85%^16,32^. Importantly, these existing tools are largely validated for use in clinical settings with only more limited self-test validation. Here, we demonstrate that the PROTECT system matches this validity using an unsupervised, self-test approach.

Validation of the system for MCI detection demonstrates good performance in identifying MCI at baseline and predicting cognitive trajectory over 24 months. This confirms that the algorithm successfully identifies MCI and that it can distinguish progressive, domain-specific neurocognitive decline from benign or reverting cognitive fluctuations. The inclusion of independent validation variables ensured that these classifications generalised to other distinct cognitive networks – such as basic psychomotor speed and complex visuospatial processing – without introducing circularity into the phenotypic validation.

Sub-typing of the MCI group into amnestic and non-amnestic phenotypes enabled more granular characterisation of the cohort. These groupings have potential clinical relevance since an amnestic MCI profile may predict a higher risk of Alzheimer’s Disease while a non-amnestic profile may indicate a non-Alzheimer trajectory trending towards synuclein or vascular dementias, so these subtypes will warrant further investigation and validation. The longitudinal analysis revealed differences in trajectory between these groups, with significant decline seen in executive function and memory in amnestic MCI which aligns to the prodromal AD phenotype. Meanwhile the non-amnestic phenotype showed a more global decline across cognitive domains.

The PROTECT system would not currently be appropriate for use as a standalone diagnostic tool since the specificity would lead to an unacceptably high level of false positive diagnoses. However, it could be used for monitoring and early triage of patients, particularly in combination with blood biomarkers to improve predictive validity. Addition of biomarkers would also facilitate improvement of specificity metrics to enable more sensitive triage of patients as part of entry into the diagnostic pathway. Recently published work indicates that the PROTECT system, in combination with capillary blood biomarkers, may facilitate identification of at-risk preclinical groups^33^. The remote nature of the system and the capillary blood technologies would allow for scaling and implementation in the community, allowing for remote self-testing of cognition in clinical pathways and trials of disease-targeted treatments. This has particular potential in pre-clinical and MCI groups where monitoring is lacking and requires a scaleable approach.

Limitations of this work must also be acknowledged. The datasets used for this study were derived from the PROTECT-UK cohort, recruited through self-referral and primary care so caution should be taken when interpreting the outcomes for a broader population. It will be important to refine the flagging algorithms in diverse independent cohorts including in underserved populations to ensure it is fully representative. Also, MCI was defined based on the FDA criteria for cognitive staging rather than a clinical diagnosis. The dementia cohort was recruited across multiple international sites and so were not precisely matched for location. As cognitive assessment was conducted in unsupervised settings and across international sites with different languages there may have been variability in consistency and approach to clinical assessments and completion of the test systems. However, this reflects the real-world nature of dementia diagnoses, and the lack of supervision aligns with the stated purpose of the system for self-testing. Seven of the eight PROTECT tests do not rely on language and so multilingualism is less of a concern than with other cognitive assessments that require verbal communication and linguistics.

Further validation is now required to refine the dementia and MCI detection algorithm in independent cohorts with a broad range of cognitive impairments and diversity characteristics, to establish prediction of clinical endpoints and correlations with brain amyloid and tau pathology using positron emission tomography and other emerging biomarkers, and to better characterise the differential trajectories of amnestic and non-amnestic MCI. Ongoing programmes of work will enable this further validation and refinement going forwards, including a large UK programme to evaluate the predictive validity of the system against clinical endpoints in people with MCI, and the potential added predictive value of blood biomarkers.

## Data Availability

All data produced in the present study are available upon reasonable request to the authors

## Acknowledgements

This paper represents independent research part-funded by the National Institute of Health Research (NIHR) Exeter Biomedical Research Centre and the NIHR HealthTech Research Centres in Brain Health and Sustainable Innovation. The views expressed are those of the authors and not necessarily those of the NIHR or the Department of Health and Social Care. This paper was also supported by the NIHR Collaboration for Leadership in Applied Health Research and Care South-West Peninsula.

## Author Contributions

Anne Corbett: Conception and design, data analysis and interpretation, manuscript drafting and reviewing; Abbie Palmer: Data acquisition and analysis, manuscript reviewing; Millie Sander: Data acquisition and analysis, manuscript reviewing; Christine Davis: Design, data acquisition, manuscript reviewing; Kim Idar Giske: data analysis and interpretation, manuscript drafting and reviewing; Kate Stych: Data acquisition, manuscript reviewing; Mary O’Leary: Data acquisition, manuscript reviewing; Chris Fox: data interpretation, manuscript reviewing; Jon Huntley: data interpretation, manuscript reviewing; Dag Aarsland: Conception and design, manuscript reviewing; Nicolas Castellano: Data acquisition, manuscript reviewing; Felipe Botero-Rodriguez: Data acquisition, manuscript reviewing; Gerard Griffieon: Data acquisition, manuscript reviewing; Mieke Nuytten: Data acquisition, manuscript reviewing; Nicholas Ashton: Data interpretation, manuscript reviewing; Hanna Huber: Data acquisition, manuscript reviewing; Jeffrey Cummings: manuscript writing and reviewing Clive Ballard: Conception and design, data analysis and interpretation, manuscript drafting and reviewing.

## Data Availability

Data are available on request from the PROTECT study following approval. Requests can be directed to

## Competing interests

Anne Corbett declares consultancy work for Novartis, Addex, Suven, Sunovion, Janssen and Acadia pharmaceutical companies and grant funding from Novo Nordisk, ReMynd, Therini Bio pharmaceutical companies. Dag Aarsland has received research support and/or honoraria from Eisai, Heptares, Eli Lilly, BioArctic, GSK, Roche Diagnostics, and discoveric bio alpha, and research support from: Muhdo Health Ltd, Daily Colors, Evonik, Sanofi and Roche Diagnostics. Gerard Griffeon is paid as a consultant for remynd and Babylon Biosciences and owns remynd warrants and shares. Nicholas Ashton has received consultancy/speaker fees from Alamar Biosciences, Bioartic, Biogen, Eli-Lilly, Neurogen Biomarking, Roche, Spear Bio, Quanterix and Vigil Neurosciences. Adam Hampshire is owner and director of Future Cognition Ltd and co-owner and co-director of H2 Cognitive Designs, which produce and provide as a service online assessment technology. Jeff Cummings has provided consultation to Acadia, Acumen, ALZpath, AnnovisBio, Artery, Axsome, Biogen, Bristol-Myers Squib, Eisai, Fosun, GAP Foundation, Hummingbird Diagnostics, IGC, Janssen, Julius Clinical, Kinoxis, Lighthouse, Lilly, Lundbeck, LSP/eqt, Merck, MoCA Cognition, Novo Nordisk, NSC Therapeutics, Optoceutics, Otsuka, ReMYND, Roche, Scottish Brain Sciences, Signant Health, Simcere, sinaptica, and T-Neuro pharmaceutical, assessment, and investment companies. JLC is is co-founder of CNS Innovations and Mangrove Therapeutics. JLC is supported by NIGMS grant P20GM109025; NIA R35AG71476; NIA R25AG083721; NINDS

RO1NS139383; Alzheimer’s Disease Drug Discovery Foundation (ADDF); Ted and Maria Quirk Endowment; Joy Chambers-Grundy Endowment. Clive Ballard has received consulting fees from Acadia pharmaceutical company, AARP, Addex pharmaceutical company, Eli Lily, Enterin pharmaceutical company, GWPharm, H.Lundbeck pharmaceutical company, Novartis pharmaceutical company, Janssen Pharmaceuticals, Johnson and Johnson pharmaceuticals, Novo Nordisk pharmaceutical company, Orion Corp pharmaceutical company, Otsuka America Pharm Inc, Sunovion Pharm. Inc, Suven pharmaceutical company, Roche pharmaceutical company, Biogen pharmaceutical company, Synexus clinical research organization and tauX pharmaceutical company and research funding from Synexus clinical research organization, Roche pharmaceutical company, Novo Nordisk pharmaceutical company and Novartis pharmaceutical company. Chris Fox has provided consultation with Tau Rx. All other authors have no competing interests to declare.

**Supplementary Table 1:**
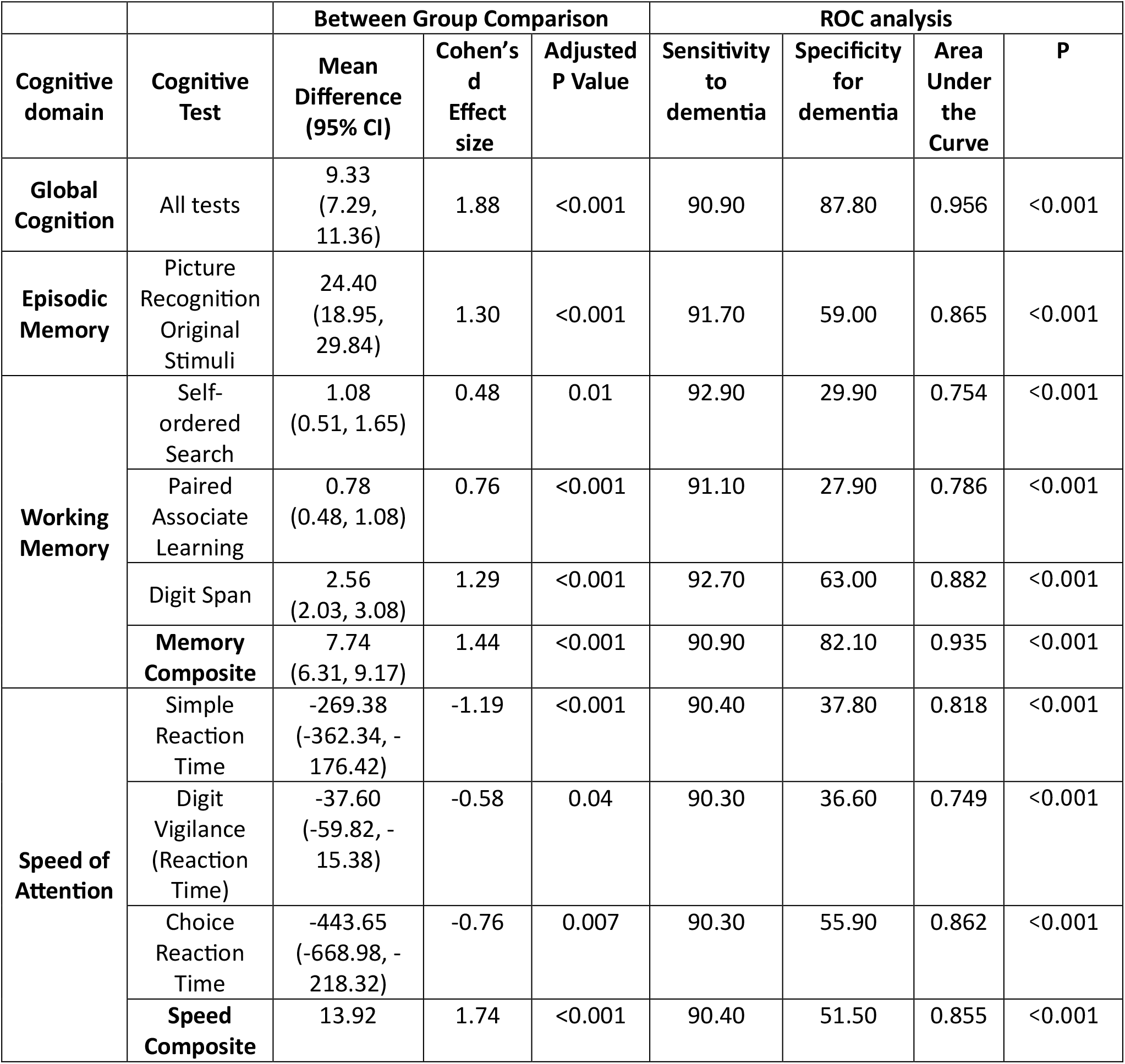
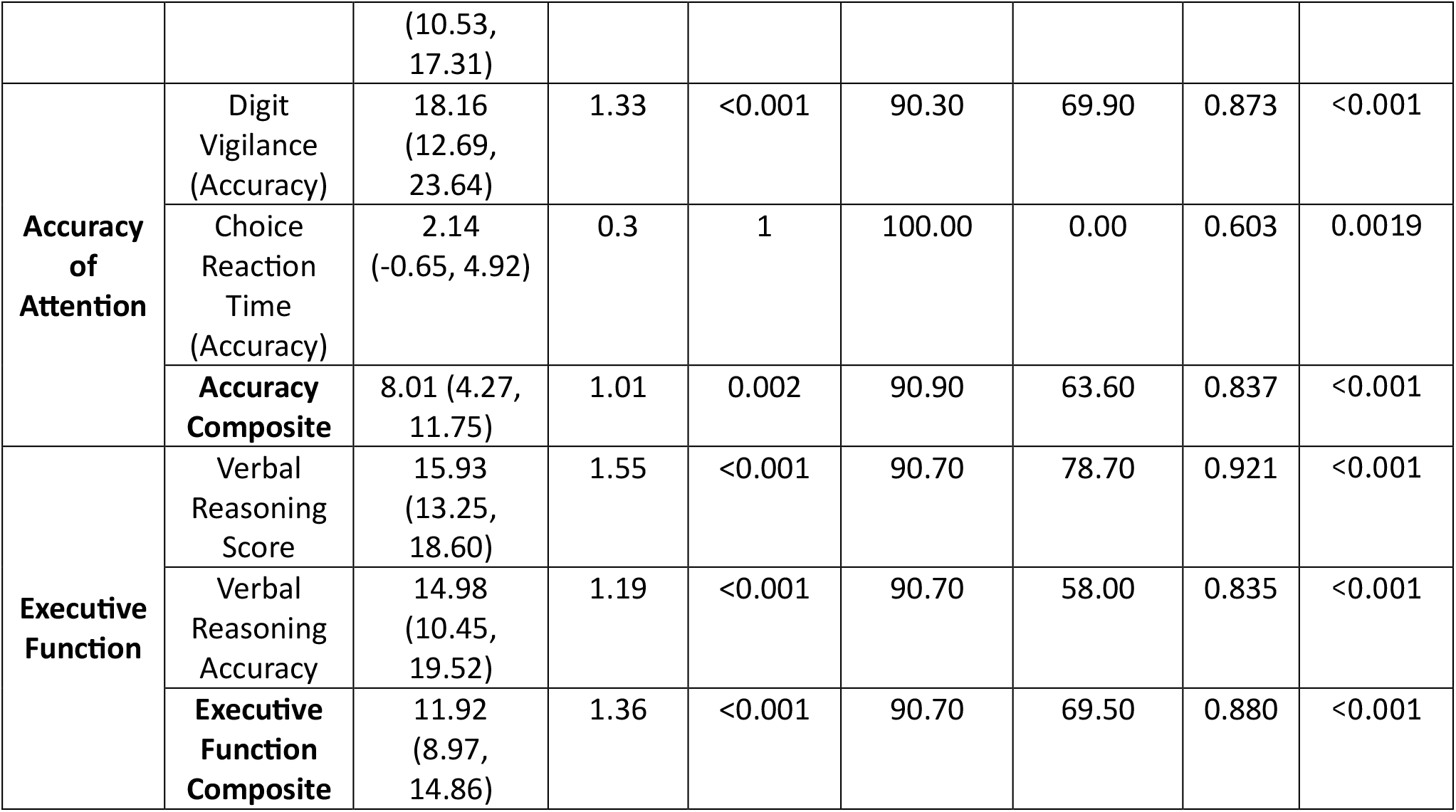
Performance of the PROTECT Cogntive Test System in between-group comparison and ROC analysis of healthy versus dementia cohorts (n=389)

## Figure Legends

**Figure 2 Legend: (A)** Box plots show the distribution of standardised scores in the composite measures of memory, speed of attention, accuracy of attention and executive function, and with the global cognition score on the PROTECT Cognitive Test System. Plots show performance in healthy (blue) and dementia (orange) cohorts. Statistical comparisons in all domains is shown in Table 2. **(B)** Distribution overlap plot showing **s**tandardised global cognition score in healthy (orange) and dementia (green) cohorts. The healthy (control) group median is shown in blue and demonstrates the significant shift in performance compared to the dementia group.

**Figure 3 Legend:** Discriminative performance of the integrated multi-domain algorithm (Global Algorithm, bold black line) and the five composite measures in differentiating the confirmed clinical dementia cohort from the healthy normative baseline (Non-MCI). The combined global algorithm yields the highest Area Under the Curve (AUC), demonstrating that multi-domain integration outperforms the diagnostic utility of any single cognitive domain in isolation. The dashed diagonal line indicates chance-level discrimination (AUC = 0.50).

**Figure 4 Legend:** Standardised Z scores at baseline for the non-MCI and MCI subtype cohorts. Solid black lines represent the normative mean. Dashed red line indicates the 1.5SD impairment threshold used for flagging of MCI.

**Figure 5 Legend:** Bars represent the ANCOVA estimated marginal differences from the healthy non-MCI controls (represented by the dashed zero-line). Subtypes are Amnestic MCI (Blue) and non-Amnestic MCI (Green). All estimates are adjusted for baseline performance, age, sex and education. Error bars represent +/-1 Standard Error.

## References

1. Alzheimer’s Research UK. Dementia Statistics UK. Alzheimer’s Research UK. Accessed 20th October, 2022.

2. Petersen RC. Mild Cognitive Impairment. Continuum (Minneap Minn). Apr 2016;22(2 Dementia):404–18. doi:10.1212/con.0000000000000313

3. Cook L, Souris, H., Isaacs, J. London memory services 2019 audit report. 2019. https://www.england.nhs.uk/london/wp-content/uploads/sites/8/2019/11/FINAL-London-memory-service-audit-2019.pdf

4. Korologou-Linden R, Kalsi J, Kafetsouli D, et al. Novel Blood-Based Biomarkers and Disease Modifying Therapies for Alzheimer’s Disease. Are We Ready for the New Era? J Prev Alzheimers Dis. 2024;11(4):897–902. doi:10.14283/jpad.2024.83

5. Laurell AAS, Venkataraman AV, Schmidt T, et al. Estimating demand for potential disease-modifying therapies for Alzheimer’s disease in the UK. Br J Psychiatry. Jun 2024;224(6):198–204. doi:10.1192/bjp.2023.166

6. Hossain MF, Husna AU, Kharel M. Use of lecanemab for the treatment of Alzheimer’s disease: A systematic review. Brain Behav. Jun 2024;14(6):e3592. doi:10.1002/brb3.3592

7. Alzheimer’s Disease: Developing Drugs for Treatment Guidance for Industry (2018).

8. Collerton J, Collerton D, Arai Y, et al. A comparison of computerized and pencil-and-paper tasks in assessing cognitive function in community-dwelling older people in the newcastle 85+pilot study. Journal of the American Geriatrics Society. Oct 2007;55(10):1630–1635. doi:10.1111/j.1532-5415.2007.01379.x

9. Chan JYC, Yau STY, Kwok TCY, Tsoi KKF. Diagnostic performance of digital cognitive tests for the identification of MCI and dementia: A systematic review. Ageing Res Rev. Dec 2021;72:101506. doi:10.1016/j.arr.2021.101506

10. Tsoy E, Zygouris S, Possin KL. Current State of Self-Administered Brief Computerized Cognitive Assessments for Detection of Cognitive Disorders in Older Adults: A Systematic Review. J Prev Alzheimers Dis. 2021;8(3):267–276. doi:10.14283/jpad.2021.11

11. Wesnes KA, Brooker H, Ballard C, McCambridge L, Stenton R, Corbett A. Utility, reliability, sensitivity and validity of an online test system designed to monitor changes in cognitive function in clinical trials. International journal of geriatric psychiatry. 2017;32(12):e83–e92.

12. Giovane MD, Giunchiglia V, Cai Z, et al. Remote cognitive tests predict neurodegenerative biomarkers in the Insight 46 cohort. Alzheimers Dement. Feb 2025;21(2):e14572. doi:10.1002/alz.14572

13. Nuytten M, Voets M, Debroux E, et al. Randomized phase 2a trial assessing a novel septin molecular glue in Alzheimer’s disease. Alzheimers Dement. Sep 2025;21(9):e70537. doi:10.1002/alz.70537

14. Hampshire A, Chatfield DA, AM MP, et al. Multivariate profile and acute-phase correlates of cognitive deficits in a COVID-19 hospitalised cohort. EClinicalMedicine. May 2022;47:101417. doi:10.1016/j.eclinm.2022.101417

15. Cubillos C, Rienzo A. Digital Cognitive Assessment Tests for Older Adults: Systematic Literature Review. JMIR Ment Health. Dec 08 2023;10:e47487. doi:10.2196/47487

16. Junkkila J, Oja S, Laine M, Karrasch M. Applicability of the CANTAB-PAL computerized memory test in identifying amnestic mild cognitive impairment and Alzheimer’s disease. Dement Geriatr Cogn Disord. 2012;34(2):83–9. doi:10.1159/000342116

17. Study. P. Platform for Research Online to investigate the genetics and cognition in ageing. http://www.protectstudy.org.uk

18. Brooker H, Williams G, Hampshire A, et al. FLAME: A computerized neuropsychological composite for trials in early dementia. Alzheimer’s & Dementia: Diagnosis, Assessment & Disease Monitoring. 2020;12(1):e12098.

19. clinicaltrials.gov. Fasudil Trial for Treatment of Early Alzheimer’s Disease (FEAD). https://clinicaltrials.gov/study/NCT06362707

20. clinicaltrials.gov. Phenserine on the Alzheimer’s Treatment Horizon, Study 1 (PATH-1). clinicaltrials.gov; 2025.

21. Corbett A, Williams G, Creese B, et al. Impact of Short-Term Computerized Cognitive Training on Cognition in Older Adults With and Without Genetic Risk of Alzheimer’s Disease: Outcomes From the START Randomized Controlled Trial. J Am Med Dir Assoc. May 2024;25(5):860–865. doi:10.1016/j.jamda.2024.03.008

22. Corbett A, Taylor R, Llewellyn D, et al. Impact of Vitamin D supplementation on cognition in adults with mild to moderate vitamin D deficiency: Outcomes from the VitaMIND randomised controlled trial. 10.1101/2024.11.21.24317708: MedRxiv; 2024.

23. ISRCTN. Validation of the FLAME Cognitive Test System in people with mild to moderate dementia. Accessed 22nd August, 2025.

24. Jack CR, Jr., Andrews SJ, Beach TG, et al. Revised criteria for the diagnosis and staging of Alzheimer’s disease. Nat Med. Aug 2024;30(8):2121–2124. doi:10.1038/s41591-024-02988-7

25. Palmqvist S, Janelidze S, Quiroz YT, et al. Discriminative Accuracy of Plasma Phospho-tau217 for Alzheimer Disease vs Other Neurodegenerative Disorders. JAMA. Aug 25 2020;324(8):772–781. doi:10.1001/jama.2020.12134

26. Ashton NJ, Brum WS, Di Molfetta G, et al. Diagnostic Accuracy of a Plasma Phosphorylated Tau 217 Immunoassay for Alzheimer Disease Pathology. JAMA Neurol. Mar 1 2024;81(3):255–263. doi:10.1001/jamaneurol.2023.5319

27. Jack CR, Jr., Andrews JS, Beach TG, et al. Revised criteria for diagnosis and staging of Alzheimer’s disease: Alzheimer’s Association Workgroup. Alzheimers Dement. Aug 2024;20(8):5143–5169. doi:10.1002/alz.13859

28. Dubois B, Villain N, Schneider L, et al. Alzheimer Disease as a Clinical-Biological Construct-An International Working Group Recommendation. JAMA Neurol. Nov 1 2024; doi:10.1001/jamaneurol.2024.3770

29. Raghavan S, Graff-Radford J, Hofrenning E, et al. Plasma NfL and GFAP for predicting VCI and related brain changes in community and clinical cohorts. Alzheimers Dement. Jun 2025;21(6):e70381. doi:10.1002/alz.70381

30. Huber H, Montoliu-Gaya L, Brum WS, et al. A minimally invasive dried blood spot biomarker test for the detection of Alzheimer’s disease pathology. Nat Med. Feb 2026;32(2):599–608. doi:10.1038/s41591-025-04080-0

31. White N, Parsons R, Collins G, Barnett A. Evidence of questionable research practices in clinical prediction models. BMC Med. Sep 04 2023;21(1):339. doi:10.1186/s12916-023-03048-6

32. White JP, Schembri A, Prenn-Gologranc C, et al. Sensitivity of Individual and Composite Test Scores from the Cogstate Brief Battery to Mild Cognitive Impairment and Dementia Due to Alzheimer’s Disease. J Alzheimers Dis. 2023;96(4):1781–1799. doi:10.3233/JAD-230352

33. Corbett A, Sander-Long, M., Ashton, N., Huber, H., Vavra, J., Braun-Wohlfahrt, L., Zetterberg, H., Montoliu-Gaya, L., Cummings, J., Bateman, F., Davis, C., Ballard, C. Alzheimer’s Disease blood biomarkers measured through remote capillary sampling correlate with cognition in older adults. Nature Communications (In Press)2026.

